# Do medical marijuana laws increase prescription stimulant use?

**DOI:** 10.1101/2023.01.29.23285169

**Authors:** Garrett D. Alexander, Luke R. Cavanah, Jessica L. Goldhirsh, Leighton Y. Huey, Brian J. Piper

## Abstract

**Background:** Chronic cannabis use can present with cognitive impairments that resemble ADHD. Our objective was to determine if medical cannabis (MC) legalization increased prescription stimulant distribution.

**Methods:** We extracted distribution of methylphenidate, amphetamine, and lisdexamfetamine from the Drug Enforcement Administration’s comprehensive database for 2006 to 2021 and compared the three-year population-corrected slopes of stimulant distribution before and after MC program implementation.

**Results:** We found a very large main effect of time (*p*≤0.001), but, contrary to the hypothesis not states’ MC sales status, on slopes of distribution (*p*=0.391). There was a significant and large interaction effect of time and MC sales status on slopes of distribution (*p*≤0.001).

**Discussion:** These findings suggest that medical cannabis program legalization may have contributed to some states having more rapid increases in Schedule II stimulant distribution rates over time.

## Introduction

There has been a significant increase in the use, and misuse, of Schedule II stimulants, which is now becoming a growing concern across the country. These stimulants have been considered the gold standard pharmacotherapy for the treatment of Attention Deficit Hyperactivity Disorder ADHD in the US (Piper, Ogden, et al., 2018). The most prescribed Schedule II stimulants used for ADHD in the US are methylphenidate, amphetamine, and lisdexamfetamine (Subcommittee on Attention-Deficit/Hyperactivity Disorder, Steering Committee on Quality Improvement and Management, 2011). Examination of comprehensive data from US Drug Enforcement Administration’s (DEA) Automation of Reports and Consolidated Orders System (ARCOS) from 2006 to 2016 identified an increase in prescription stimulant use over time. Although the US population increased only 8%, between 2006 and 2016 there was a 2.5-fold increase in amphetamine use while lisdexamfetamine steadily increased each year since its 2007 approval (Piper, Ogden, et al., 2018). In addition, the US was responsible for over 80% of the global consumption of methylphenidate (International Narcotics Control Board et al., 2016). There was an uptick of amphetamine-type stimulants and long-acting type stimulants distribution from 5.6 to 6.1 prescriptions per 100 persons from 2014 to 2019 (Board et al., 2020).

Moreover, there are several lines of evidence that support the idea that there is an association between marijuana use and the increase in stimulant use (Haffajee & Heins, 2021). The use of marijuana can impair cognitive functioning which can mimic symptoms of ADHD. Because of this, there is emerging evidence that illustrates a link between marijuana and stimulant usage (American Psychiatric Association, 2013). Studies have shown that marijuana worsens executive function and working memory, which are the areas patients with ADHD are deficient. Studies have also shown that marijuana can exacerbate neurocognitive function, especially for people who have ADHD. Therefore, marijuana can have an additive effect for those with ADHD, worsening their ability to focus and get tasks done efficiently (Mitchell, et al., 2016). Also, those who used stimulants were more than four times as likely to use cannabis (Haffajee & Heins, 2021). Almost three-quarters (74%) of states in the US have approved MC (*State Medical Cannabis Laws*, n.d.).

The purpose of this study was to analyze data from ARCOS, which reports distribution of stimulants methylphenidate, amphetamine, and lisdexamfetamine (Piper, Ogden, et al., 2018) and determine if there was a significant difference in stimulant use pre-versus post-MC implementation. We hypothesized that distribution of stimulants would significantly increase after legalization for states with (MC+) but not without (MC−).

## Methods

### Procedures

Data was extracted from the DEA’s ARCOS, a comprehensive national database containing a yearly updated report of retail drug distribution from manufacturers and distributors (Drug Enforcement Administration, Diversion Control Division, 2021). This database has been frequently used in prior pharmacoepidemiology reports (Atluri et al., 2014; Bokhari et al., 2005; Collins et al., 2019; Davis et al., 2020; Pashmineh Azar et al., 2020; Piper et al., 2020; Simpson et al., 2019; Vaddadi et al., 2021). ARCOS was validated by examining the total weight of oxycodone in this database relative to that reported in a Prescription Monitoring Program which revealed a high correlation (*r* = 0.99) (Piper, Shah, et al., 2018). Extracted data included total grams of stimulant use from 2006 to 2021. Three Schedule II stimulants were examined: amphetamine, methylphenidate, and lisdexamfetamine.

We examined the change in stimulant distribution of these Schedule II stimulants before and after MC program implementation. To evaluate the changes in number of patients utilizing the Schedule II stimulants of interest, we used the median estimated Daily Dosage per person (mg/person/day) for each stimulant (Vaddadi et al., 2021). We defined the start dates of MC programs as the quarter where MC sales first began in the state. These data were found on either the state’s MC program website or from (Kaufman et al., 2021). The research was deemed exempt by the Geisinger IRB.

### Data Analysis

Total weight was calculated for each drug per quarter per year for the fifty states and the District of Columbia. A population-corrected index for each stimulant was calculated as the weight divided by the municipality’s population as estimated by the US American Community Survey (ACS). Stimulant use was compared from 2006-2021 for states with versus without MC (+ versus -). If the assumption of variance was not met (*p*<0.10), a separate variance t-test was completed. Subsequently, an interrupted time series was performed. Slopes for individual and total stimulant use from three years before the implementation of MC (pre-) and three years after (post-) were calculated for MC+ and MC−states. The quarter of implementation was excluded as a transitional period. The mean MC program implementation date (first quarter of 2016) was used as the interruption point for the comparison group. A two-way mixed ANOVA was used to determine the effect of MC time relative to MC implementation (pre, post) and MC legal sales status (+, -) on slopes of state prescription stimulant distribution, which was reported in units of grams per million people per year for amphetamine, lisdexamfetamine, and methylphenidate, and daily doses per hundred million people per year for the total. Effect size was calculated as partial eta-squared (η_*p*_^2^). Identical analyses were conducted using slopes two years before and two years after MC implementation. Data was analyzed using Excel and SPSS and figures were made with Heatmapper (Babicki et al., 2016) and Graph Pad Prism

## Results

### Time

We found the anticipated very large effects of time on slopes of state amphetamine, lisdexamfetamine, methylphenidate, and total stimulant use. Contrary to the hypothesis, Figure 1 shows that, for MC+ states, the slopes of amphetamine, lisdexamfetamine, methylphenidate, and their sum were each significantly smaller after implementation of MC programs. Similarly, Figure 1 shows that, for MC−states, the slopes of amphetamine, lisdexamfetamine, methylphenidate, and their sum were each significantly smaller after the implementation time of MC programs.

**Figure 1:**
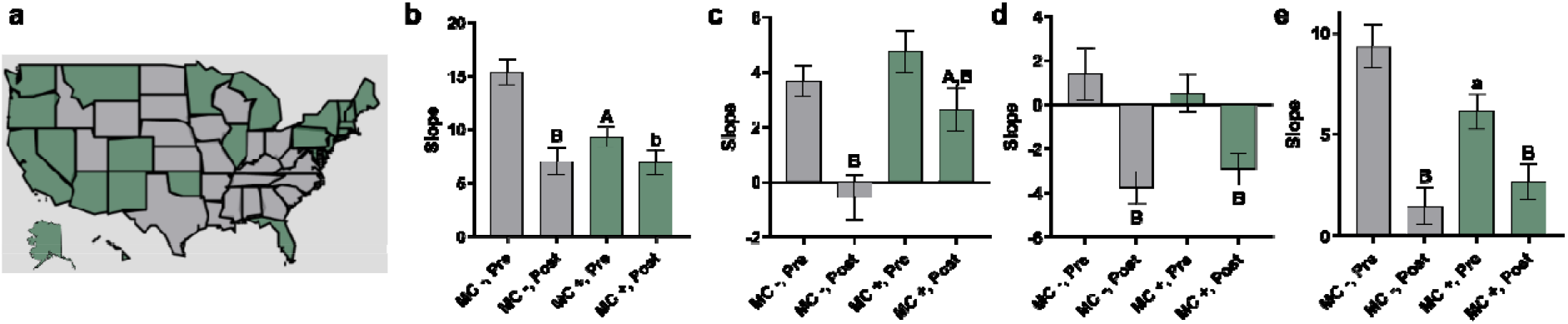
Slopes of Stimulant Distribution Rates as Function of Medical Cannabis Sales. *Note*. **a** Heat map of the US by medical cannabis (MC) sales status (green = sales; grey = no sales). ADHD Stimulant (**b** amphetamine, **c** lisdexamfetamine, **d** methylphenidate, **e** sum) use in the US as reported by the DEA’s ARCOS from 2006-2021 as a function of presence (+) or absence (−) of state MC program implementation. Slopes for stimulants distribution rates three years (before) and three years after (post) state MC implementation (^a^p<0.05 versus corresponding MC−, ^b^p<0.05 versus corresponding pre, ^A^p<0.001 versus corresponding MC−, ^B^p<0.001 versus corresponding pre). For individual stimulants, slopes are in units of grams per million people per year; for aggregate stimulant slopes, slopes are in units of daily doses per hundred million people per year. Error bars denote standard errors of the mean.

### Cannabis Status

As hypothesized, we found moderate effects of MC status on slopes of amphetamine and lisdexamfetamine. No significant effect of MC status was found on slopes of methylphenidate or the total. Not only were significant differences found between pre- and post-conditions, but also between MC+ and the respective MC−conditions: for the pre-MC period, slopes for amphetamine and the total were significantly smaller in states that went on to implement MC compared to states that did not. No significant differences existed between the MC+ and MC−states in the pre-conditions for lisdexamfetamine or methylphenidate. Additionally, for the post-MC period, the slope for lisdexamfetamine was significantly larger in MC+ compared to MC−states. No other significant differences noted between post conditions.

### Interaction of Time and Cannabis Status

We found very large interaction effects of time and MC status on slopes of amphetamine, lisdexamfetamine, and total. No statistically significant interaction effect of time and MC status was observed for methylphenidate. No statistically significant interaction effect of time and MC status was observed for any of the stimulants or the total.

Supplemental analyses using two-year regression slopes, were conducted, and reveal some differences (Supplemental Figure 1).

## Discussion

The purpose of this study was to examine the effects of MC program legalization on Schedule II stimulant distribution. Based on the results, there was a significant decrease in slope values during the pre-MC legalization period for states that subsequently legalized MC compared to non-MC states. However, there was not a consistent significant difference for pre versus post MC program implementation across all three stimulants we studied: amphetamine, methylphenidate, and lisdexamfetamine. This finding is contrary to the hypothesis that MC legalization has had a pronounced influence on the increase in stimulant use across the US in the past two decades.

Although this seems contrary to what was expected, there was some evidence that supported that MC legalization does have an impact on the increase in stimulant distribution. Based on the total stimulant three-year slopes bar graph (Figure 1), there was a significant difference between pre- and post-MC program implementation. This shows the potential for a connection between MC legalization and the increase in stimulants that has been observed throughout the US. Additionally, when looking at the total stimulant slope values (Figure 1), there is a greater decrease in slope values for MC− states versus MC + states when comparing slopes pre against post medical cannabis program implementation. Consequently, this finding lends support to MC programs having an influence on stimulant usage because there was a smaller decrease in slope values when comparing pre versus post MC program implementation. This trend illustrates that MC states are more resistant to a negative decrease in slope values which could potentially mean that MC implementation played a role in the increase in Schedule II stimulants prescriptions.

However, for the amphetamine, there was no significant difference in the two- and three-year slopes for pre versus post MC + states. On the other hand, lisdexamfetamine, methylphenidate, and total stimulants illustrated a trend that showed no significance for the two-year slopes but a significant difference for the three-year slopes. Therefore, this data, with an exception to amphetamine, generally supports the idea that there was a significant difference for MC + states when comparing pre-against post-MC implementation. The longer timeframe for the three-year slopes could contribute to why there was a general significant difference seen compared to the two-year slopes where there is less time to observe the effects on MC across the states.

Moreover, there are other variables that impact the upward pattern seen in stimulant prescriptions. For adult ADHD, the number of criteria that patients must meet to qualify for the condition was altered from 6 to 5 criteria needed for diagnosis in 2013 (Epstein & Loren, 2013). ADHD testing for children, especially in low-income areas, has increased dramatically, which research has shown has been influenced by the establishment of the No Child Left Behind (NCLB) Act of 2001, resulting in an increase in ADHD diagnosis amongst the youth (Fulton et al., 2015). The idea is that the establishment of the NCLB Act created trepidation in many school districts of the potential for loss of school funds for poor performing children on standardized tests. Thus, schools increased testing for ADHD for struggling students which resulted in increased stimulants prescribed for students to help improve test scores. This is supported by data that showed a doubling of ADHD diagnosis in low-income areas across states from 2003 to 2007 in places that were introduced to the increase in ADHD testing as part of the NCLB program. Therefore, the combined effect of increased inclusion of ADHD diagnostic criteria for adults in the DSM-5 and increased testing for ADHD amongst children could contribute to the increase in ADHD diagnoses, subsequently resulting in an increase in Scheduled II stimulant prescriptions.

Additionally, the use of stimulants such as lisdexamfetamine for Binge Eating Disorder has likely contributed to the increase as well (Guerdjikova et al., 2016). As a prescription cannabinoid agonist has been approved for cachexia in HIV patients, and a cannabinoid antagonist was employed briefly in Europe to treat obesity (Marco et al., 2012), future pharmacoepidiological investigation could examine whether cannabis laws impact state-level differences in body weight. Moreover, a limitation of this study that could have an influence on stimulant use is the impact of recreational cannabis legalization. Recreational marijuana use has increased access to cannabis to the general population which cannot be accounted for in this study since the focus was on MC. Another caveat is that lisdexamfetamine did not start to be prescribed until 2007 in the US (Elbe et al., 2010). This means that the implementation for prescription use of this drug may vary widely across the states since it is newer compared to the other two drugs studied. This might also explain why the slopes for lisdexamfetamine were different then the slope for the other stimulants examined. The US accounted for less than five-percent of the world’s population but over four-fifths (83.1%) of the global volume of ADHD medications (Scheffler et al., 2007). This finding should be verified in other countries that have had substantial increases in prescription stimulants (amphetamine, lisdexamfetamine, methylphenidate) and that have increased their availability of cannabis. Additionally, examination of the potential impact of cannabis legalization on ADHD diagnoses and prescription rates of non-stimulant pharmacologic treatments of ADHD may be an interesting future pursuit.

Fortunately, when observing the total stimulant graph, there is a recognizable pattern seen when comparing pre against post MC program implementation, which supports the hypothesis that MC programs do impact stimulant use to an extent. Furthermore, there are various factors that are influencing the increase in Schedule II stimulant usage, which indicates further investigation and statistical analyses are necessary to develop a more concrete understanding of the effects of MC implementation on the increase in stimulant use in the US over the last two decades.

In conclusion, the dynamic changes in MC policy in the United States may have intended, and perhaps unintended, consequences. This investigation partially supports the hypothesis that MC implementation contributes to modulating the state-level differences in the population level distribution of prescription stimulants. Further research at a patient level using electronic medical records may be warranted.

## Supporting information

Supplemental Raw Data

## Data Availability

All data produced in the present study are available upon reasonable request to the authors and available as supplemental material on medrxiv.org

https://www.deadiversion.usdoj.gov/arcos/retail_drug_summary/index.html

