## Supplemental Tables and Figures for "Do medical marijuana laws increase prescription stimulant use?"

**Supplemental Figure 1**

*Slopes of Stimulant Distribution Rates as Function of Medical Cannabis Sales (Two Years Pre, Two Years Post)*

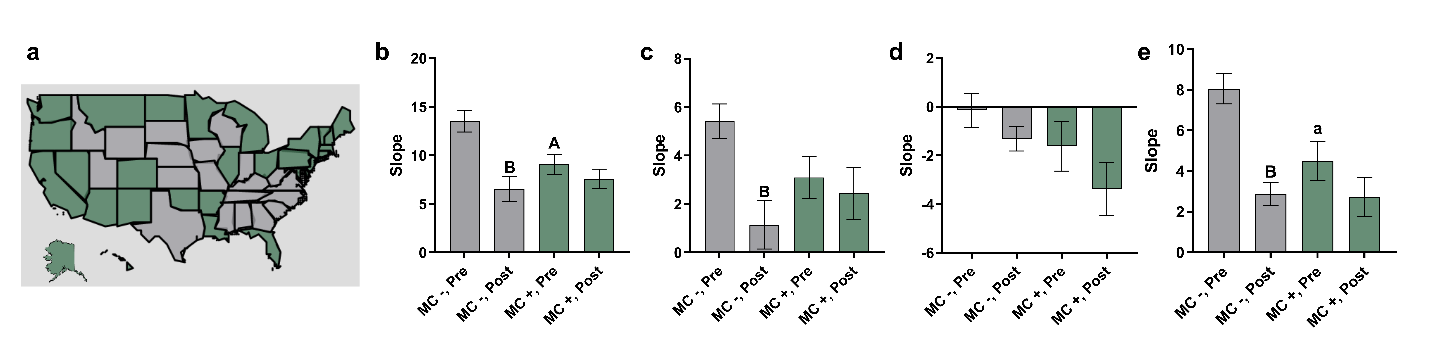
*Note.* **a** Heat map of the US by medical cannabis (MC) sales status (green = sales; grey = no sales). ADHD Stimulant (**b** amphetamine, **c** lisdexamfetamine, **d** methylphenidate, **e** sum) use in the US as reported by the DEA’s ARCOS from 2006-2021 as a function of presence (+) or absence (-) of state MC program implementation. Slopes for stimulants distribution rates two years (before) and two years after (post) state MC implementation (^a^p<0.05 versus corresponding MC -, ^b^p<0.05 versus corresponding pre, ^A^p<0.001 versus corresponding MC -, ^B^p<0.001 versus corresponding pre). For individual stimulants, slopes are in units of grams per million people per year; for aggregate stimulant slopes, slopes are in units of daily doses per hundred million people per year. Error bars denote standard errors of the mean.

**Supplemental Table 1**

*Medical cannabis states included in this report and the year of first medical sales of cannabis, and the source of the information.*

| State | Year | Source |
| --- | --- | --- |
| Alaska | 2016 | <https://www.mpp.org/issues/medical-marijuana/state-by-state-medical-marijuana-laws/medical-marijuana-program-implementation-timeline/> |
| Arizona | 2012 | <https://azcapitoltimes.com/welcome-ad/?retUrl=/news/2012/12/06/first-medical-marijuana-dispensary-in-arizona-starts-selling-pot/> |
| Arkansas | 2019 | <https://sanctuarywellnessinstitute.com/cannabis/arkansas/medical-marijuanas-law.php> |
| California | 2018 | <https://www.mpp.org/issues/medical-marijuana/state-by-state-medical-marijuana-laws/medical-marijuana-program-implementation-timeline/> |
| Colorado | 2014 | [https://www.mpp.org/issues/medical-marijuana/state-by-state-medical-marijuana-laws/medical-marijuana-program-implementation-timeline//](https://www.mpp.org/issues/medical-marijuana/state-by-state-medical-marijuana-laws/medical-marijuana-program-implementation-timeline/) |
| Connecticut | 2014 | <https://www.courant.com/business/hc-medical-marijuana-sales-begin-connecticut-20140922-story.html> |
| Delaware | 2015 | <https://dhss.delaware.gov/dhss/dph/hsp/files/mmpannrpt2017.pdf> |
| District of Columbia | 2013 | <https://www.washingtonpost.com/local/dc-politics/dc-records-its-first-legal-pot-deal-in-at-least-75-years/2013/07/29/17521b42-f889-/> |
| Florida | 2016 | <https://www.mpp.org/issues/medical-marijuana/state-by-state-medical-marijuana-laws/medical-marijuana-program-implementation-timeline/> |
| Hawaii | 2017 | <https://www.mpp.org/issues/medical-marijuana/state-by-state-medical-marijuana-laws/medical-marijuana-program-implementation-timeline/> |
| Illinois | 2015 | <https://www.chicagotribune.com/news/ct-illinois-medical-marijuana-list-met-20151113-story.html> |
| Louisiana | 2019 | <https://www.mpp.org/issues/medical-marijuana/state-by-state-medical-marijuana-laws/medical-marijuana-program-implementation-timeline/> |
| Maine | 2011 | <https://www.pressherald.com/2015/03/02/medical-pot-sales-tax-income-growing-in-maine/> |
| Maryland | 2017 | <https://www.mpp.org/issues/medical-marijuana/state-by-state-medical-marijuana-laws/medical-marijuana-program-implementation-timeline/> |
| Massachusetts | 2015 | <https://www.wbur.org/news/2015/10/08/medical-marijuana-sales-massachusetts> |
| Michigan | 2018 | [https://www.mpp.org/issues/medical-marijuana/state-by-state-medical-marijuana-laws/medical-marijuana-program-implementation-timeline//](https://www.mpp.org/issues/medical-marijuana/state-by-state-medical-marijuana-laws/medical-marijuana-program-implementation-timeline/) |
| Minnesota | 2015 | <https://www.twincities.com/2017/05/15/minnesotas-medical-marijuana-providers-have-lost-a-combined-11-million/> |
| Missouri | 2020 | [https://www.mpp.org/issues/medical-marijuana/state-by-state-medical-marijuana-laws/medical-marijuana-program-implementation-timeline//](https://www.mpp.org/issues/medical-marijuana/state-by-state-medical-marijuana-laws/medical-marijuana-program-implementation-timeline/) |
| Montana | 2009 | <https://www.mpp.org/states/montana/montanas-medical-marijuana-laws-history/> |
| Nevada | 2015 | <https://www.mpp.org/issues/medical-marijuana/state-by-state-medical-marijuana-laws/medical-marijuana-program-implementation-timeline/> |
| New Hampshire | 2016 | <https://www.mpp.org/issues/medical-marijuana/state-by-state-medical-marijuana-laws/medical-marijuana-program-implementation-timeline/> |
| New Jersey | 2012 | <https://www.google.com/url?sa=D&q=https://www.nydailynews.com/news/national/dispensary-medical-pot-wins-permit-n-article-1.1185118&ust=1656433080000000&usg=AOvVaw3s2Qxsmo4SINYHSrLINIsB&hl=en> |
| New Mexico | 2010 | <https://www.mpp.org/issues/medical-marijuana/state-by-state-medical-marijuana-laws/medical-marijuana-program-implementation-timeline/> |
| New York | 2016 | <https://www.mpp.org/issues/medical-marijuana/state-by-state-medical-marijuana-laws/medical-marijuana-program-implementation-timeline/> |
| North Dakota | 2019 | <https://www.mpp.org/issues/medical-marijuana/state-by-state-medical-marijuana-laws/medical-marijuana-program-implementation-timeline/> |
| Ohio | 2019 | <https://www.mpp.org/issues/medical-marijuana/state-by-state-medical-marijuana-laws/medical-marijuana-program-implementation-timeline/> |
| Oklahoma | 2018 | [https://www.mpp.org/issues/medical-marijuana/state-by-state-medical-marijuana-laws/medical-marijuana-program-implementation-timeline//](https://www.mpp.org/issues/medical-marijuana/state-by-state-medical-marijuana-laws/medical-marijuana-program-implementation-timeline/) |
| Oregon | 2014 | <https://www.mpp.org/issues/medical-marijuana/state-by-state-medical-marijuana-laws/medical-marijuana-program-implementation-timeline/> |
| Pennsylvania | 2018 | <https://www.mpp.org/issues/medical-marijuana/state-by-state-medical-marijuana-laws/medical-marijuana-program-implementation-timeline/> |
| Rhode Island | 2013 | <https://www.bostonglobe.com/metro/2013/04/19/first-medical-marijuana-dispensary-opens/ChkjssrdG5Sv9GWOr6wcVO/story.html> |
| Utah | 2020 | <https://www.mpp.org/issues/medical-marijuana/state-by-state-medical-marijuana-laws/medical-marijuana-program-implementation-timeline/> |
| Vermont | 2013 | <https://mjbizdaily.com/vermont-becomes-12th-state-with-medical-marijuana-dispensaries-creating-2-million-market/> |
| Virginia | 2020 | <https://www.mpp.org/issues/medical-marijuana/state-by-state-medical-marijuana-laws/medical-marijuana-program-implementation-timeline/> |
| Washington |  | <https://www.mpp.org/issues/medical-marijuana/state-by-state-medical-marijuana-laws/medical-marijuana-program-implementation-timeline/> |

**Supplemental Table 2**

*Descriptive Statistics of Stimulant Two-Year Distribution Slopes Before and After Medical Cannabis Implementation*

|  | MC - (*N* = 20) | | | | MC + (*N* = 31) | | | | Total (*N* = 51) | | | |
| --- | --- | --- | --- | --- | --- | --- | --- | --- | --- | --- | --- | --- |
|  | Pre | | Post | | Pre | | Post | | Pre | | Post | |
|  | *M* | *SD* | *M* | *SD* | *M* | *SD* | *M* | *SD* | *M* | *SD* | *M* | *SD* |
| Total^a^ | 8.05 | 3.35 | 2.88 | 2.51 | 4.50 | 5.38 | 2.71 | 5.37 | 5.90 | 4.97 | 2.78 | 4.44 |
| Amphetamine^b^ | 13.54 | 5.03 | 6.51 | 5.70 | 9.08 | 5.64 | 7.58 | 5.67 | 10.83 | 5.79 | 7.16 | 5.65 |
| Lisdexamfetamine^b^ | 5.41 | 3.22 | 1.14 | 4.45 | 3.10 | 4.75 | 2.45 | 5.96 | 4.01 | 4.33 | 1.93 | 5.41 |
| Methylphenidate^b^ | -0.14 | 3.12 | -1.32 | 2.24 | -1.62 | 5.71 | -3.37 | 6.09 | -1.04 | 4.88 | -2.56 | 5.02 |

^a^ units of daily doses per hundred million people per year

^b^ units are in units of grams per million people per year

**Supplemental Table 3**

*Results of Independent t-test Examining Effect of States’ Medical Cannabis Sales Status on Two-Year Slopes of State Stimulant Distribution Rates*

|  | Pre | | | Post | | |
| --- | --- | --- | --- | --- | --- | --- |
| Drug | *t*(49) | *p* | Cohen's *d* | *t*(49) | *p* | Cohen's *d* |
| Total | 2.63 | 0.011* | 0.755 | 0.13 | 0.897 | 0.037 |
| Amphetamine | 2.87 | 0.006** | 0.824 | -0.65 | 0.516 | 0.188 |
| Lisdexamfetamine | 1.91 | 0.062 | 0.548 | -0.84 | 0.405 | 0.241 |
| Methylphenidate | 1.06 | 0.294 | 0.304 | 1.44 | 0.155 | 0.414 |

*p<0.05. **p<0.01

**Supplemental Table 4**

*Results of Paired t-test Examining Effect of Time Relative to Medical Cannabis Sales Status on Two-Year Slopes of State Stimulant Distribution Rates*

|  | MC + | | | MC - | | |
| --- | --- | --- | --- | --- | --- | --- |
| Drug | *t*(30) | *p* | Cohen's *d* | *t*(19) | *p* | Cohen's *d* |
| Total | 1.45 | 0.159 | 0.260 | 10.40 | <0.001** | 2.326 |
| Amphetamine | 1.33 | 0.193 | 0.239 | 6.21 | <0.001** | 1.388 |
| Lisdexamfetamine | 1.04 | 0.306 | 0.187 | 4.14 | <0.001** | 0.926 |
| Methylphenidate | 1.17 | 0.253 | 0.210 | 1.18 | 0.254 | 0.263 |

*p<0.05. **p<0.01

**Supplemental Table 5**

*Results of Mixed Methods ANOVA Examining Effect of Time Relative to Medical Cannabis Sales Status and MC Sales Status on Two-year Slopes of State Prescription Stimulant Distribution Rates*

|  | Time | | | MC Status | | | Interaction | | |
| --- | --- | --- | --- | --- | --- | --- | --- | --- | --- |
|  | *F*(1) | *P* | *η_p_*^2^ (%) | *F*(1) | *p* | *η_p_*^2^ (%) | *F*(1) | *p* | *η_p_*^2^ (%) |
| Total | 18.99 | <0.001** | 28 | 3.15 | 0.082 | 6 | 4.48 | 0.040* | 8.4 |
| Amphetamine | 25.85 | 0.002** | 35 | 1.57 | 0.216 | 3.1 | 10.82 | 0.002** | 18.0 |
| Lisdexamfetamine | 18.79 | 0.003** | 28 | 0.16 | 0.694 | 0.3 | 10.15 | 0.003** | 17.0 |
| Methylphenidate | 2.06 | 0.157 | 4 | 3.29 | 0.076 | 6.3 | 0.08 | 0.780 | 0.2 |

*p<0.05. **p<0.01

**Supplemental Table 6**

*Descriptive Statistics of Stimulant Three-Year Distribution Slopes Before and After Medical Cannabis Implementation*

|  | MC - (*N* = 24) | | | | MC + (*N* = 27) | | | | Total (*N* = 51) | | | |
| --- | --- | --- | --- | --- | --- | --- | --- | --- | --- | --- | --- | --- |
|  | Pre | | Post | | Pre | | Post | | Pre | | Post | |
|  | *M* | *SD* | *M* | *SD* | *M* | *SD* | *M* | *SD* | *M* | *SD* | *M* | *SD* |
| Total^a^ | 9.33 | 5.22 | 1.47 | 4.48 | 6.11 | 4.48 | 2.67 | 4.62 | 7.63 | 5.06 | 2.10 | 4.55 |
| Amphetamine^b^ | 15.44 | 5.90 | 6.99 | 6.27 | 9.31 | 5.12 | 6.93 | 6.06 | 12.19 | 6.26 | 6.96 | 6.10 |
| Lisdexamfetamine^b^ | 3.67 | 2.73 | -0.55 | 3.99 | 4.76 | 3.95 | 2.65 | 4.08 | 4.25 | 3.44 | 1.14 | 4.31 |
| Methylphenidate^b^ | 1.39 | 5.91 | -3.77 | 3.51 | 0.53 | 4.46 | -2.92 | 3.60 | 0.94 | 5.16 | -3.32 | 3.55 |

^a^ units of daily doses per hundred million people per year

^b^ units are in units of grams per million people per year

**Supplemental Table 7**

*Results of Independent t-test Examining Effect of States’ Medical Cannabis Sales Status on Three-Year Slopes of State Stimulant Distribution Rates*

|  | Pre | | | Post | | |
| --- | --- | --- | --- | --- | --- | --- |
| Drug | *t*(49) | *p* | Cohen's *d* | *t*(49) | *p* | Cohen's *d* |
| Total | 2.37 | 0.022* | 0.665 | -0.93 | 0.355 | 0.262 |
| Amphetamine | 3.97 | <0.001** | 1.114 | 0.04 | 0.972 | 0.010 |
| Lisdexamfetamine | -1.14 | 0.262 | 0.319 | -2.83 | 0.007** | 0.793 |
| Methylphenidate | 0.59 | 0.556 | 0.166 | -0.85 | 0.401 | 0.238 |

*p<0.05. **p<0.01

**Supplemental Table 8**

*Results of Paired t-test Examining Effect of Time Relative to Medical Cannabis Sales Status on Three-Year Slopes of State Stimulant Distribution Rates*

|  | MC + | | | MC - | | |
| --- | --- | --- | --- | --- | --- | --- |
|  | *t*(26) | *p* | Cohen's *d* | *t*(23) | *p* | Cohen's *d* |
| Total | 3.91 | <0.001** | 0.753 | 9.73 | <0.001** | 1.986 |
| Amphetamine | 2.47 | 0.020* | 0.475 | 7.31 | <0.001** | 1.492 |
| Lisdexamfetamine | 4.16 | <0.001** | 0.800 | 4.99 | <0.001** | 1.019 |
| Methylphenidate | 3.87 | <0.001** | 0.744 | 4.94 | <0.001** | 1.009 |

*p<0.05. **p<0.01

**Supplemental Table 9**

*Results of Mixed-Methods ANOVA Examining Effects of Time and Medical Cannabis Sales Status on Three-Year Slopes of State Prescription Stimulant*

|  | Time | | | MC Status | | | Interaction | | |
| --- | --- | --- | --- | --- | --- | --- | --- | --- | --- |
|  | *F*(1) | *p* | *η_p_*^2^ | *F*(1) | *p* | *η_p_*^2^ (%) | *F*(1) | *p* | *η_p_*^2^ (%) |
| Total | 87.96 | <0.001** | 64.0 | 0.75 | 0.391 | 1.5 | 13.42 | < 0.001** | 22.0 |
| Amphetamine | 52.51 | 0.002** | 52.0 | 4.50 | 0.039* | 8.5 | 16.47 | 0.002** | 25.0 |
| Lisdexamfetamine | 43.37 | 0.003** | 47.0 | 5.28 | 0.026* | 9.7 | 4.79 | 0.033* | 8.9 |
| Methylphenidate | 39.80 | 0.004** | 45.0 | 0 | 0.994 | 0 | 1.57 | 0.216 | 3.1 |

*p<0.05. **p<0.01
